# Age Pattern of Premature Mortality under varying scenarios of COVID-19 Infection in India

**DOI:** 10.1101/2020.06.11.20128587

**Authors:** Sanjay K Mohanty, Umakanta Sahoo, Udaya S Mishra, Manisha Dubey

## Abstract

**Background:** India is vulnerable to community infection of COVID-19 due to crowded and poor living condition, high density, slums in urban areas and poor health care system. The number of COVID 19 infection has crossed 300,000 with over 7,500 deaths despite a prolonged period of lock down and restrictions in public spaces. Given the likely scale and magnitude of this pandemic, it is important to understand its impact on the age pattern of mortality under varying scenarios.

**Objective:** The main objective of this paper is to understand the age pattern of mortality under varying scenarios of community infection.

**Data and Methods:** Data from the Sample Registration System (SRS), covidi19india.org and country specific data from worldmeter is used in the analyses. Descriptive statistics, case-fatality ratio, case fatality ratio with 14 days delay, abridged life table,years of potential life lost (YPLL) and disability adjusted life years (DALY) is used.

**Results:** The case fatality ratio (CFR) with 14 days delay for India is at least twice higher (8.0) than CFR of 3.4. Considering 8% mortality rate and varying scenario of community infection by 0.5%, 1% and 2%, India’s life expectancy will reduce by 0.8, 1.5 and 3.0 years and potential life years lost by 12.1 million, 24.3 million and 48.6 million years respectively. A community infection of 0.5% may result in DALY by 6.2 per 1000 population. Major share of PYLL and DALY is accounted by the working ages.

**Conclusion:** COVID-19 has a visible impact on mortality with loss of productive life years in working ages. Sustained effort at containing the transmission at each administrative unit is recommended to arrest mortality owing to COVID-19 pandemic.

**What is known?:** The case fatality rate associated with COVID-19 is low in India compared to many other countries. The mortality level is higher among elderly and people with co-morbidity.

**Contribution:** The case fatality ratio is illusive in the sense that the same with 14 days delay for India is at least twice higher (8.0). The COVID-19 attributable mortality has the potential to reduce the longevity of the population. Unlike developed countries, about half of the COVID-19 attributable mortality would be in the working age group of 45-64 years. With any level of community infection, the years of potential life lost (YPLL) and disability adjusted life years (DALY) world be highest in the working age group (45-64 years).

## Introduction

With additional 350 thousand deaths in last five months, the global share of COVID-19 attributable death has increased by 0.05% (https://population.un.org/wpp/DataQuery/). The first case surfacing in Wuhan city of China in December, the estimated number of confirmed COVID-19 cases has crossed 5 million mark by 20^th^ May 2020 (https://www.worldometers.info/). Despite this brief duration of this outbreak, the epidemic has engulfed the entire world. The COVID-19 virus is highly contagious with a longer survival duration affects the vital organs including the respiratory system. It has altered the public health priority globally and nationally and posed a challenge to medical science.

The outbreak of COVID-19 has led to a health emergency and economic crisis worldwide. The global recession following COVID-19 epidemic is projected to be worst ever in human history. In developing countries, the hard hit are the poor, migrant labour, small and petty shopkeeper; who have lost their livelihood (Bhagat et al. 2020). Many adverse economic effects are yet to surface coming days. The adverse impact of COVID-19 on human health includes raised the levels of mortality and morbidity level, treatment complications, increased health spending and reduced spending on non-health goods and services. The case fatality rate from the epidemic is high among elderly and critically ill patients such as cancer patient, hypertensive, diabetic and patients with heart disease and for those with co-morbidity (Chow et al 2020; Bonow et al. 2020; Lippi et al. 2020; Pal & Bhadada 2020; Guan et al. 2020)

The medical know-how on treatment of COVID-19 is limited and there is no effective vaccine to prevent the deadly virus. An effective vaccine for the COVID-19 is set to be in place between six month and two years from now. Besides, the health infrastructure across countries is under severe strain for treatment of COVID-19. Countries such as Italy, United States and Spain with reasonably well-developed health care system in the world could not limit the human loss in wake of COVID-19 epidemic. The Institute for Health Metric has predicted that deaths from COVID-19 in United States would lead to scarcity of medical beds and equipment (Murray et al. 2020).

The first empirical study of Diamond Princes cruise ship of Japan placed the COVID-19 infection fatality rate at 1.3%, corrected case fatality ratio at 2.6% for all age group and 13% among the older aged 70+ (Russell et al. 2020). The mortality rate due to COVID-19 and its bearing on life expectancy is higher in countries with higher proportion of adults and elderly (Lai et al. 2020). The overall crude fatality rate from COVID-19 in China was greater among the elderly aged 60 and above as well as males (Verity et al. 2020; Dudley et al. 2020). The COVID 19 fatality is responsive to population aging and health care resources (Dilcher et al. 2020; Ji et al. 2020). Italy had a larger share of confirmed COVID-19 infection among elderly and those with multiple comorbidities (Onder et al. 2020). Myocardial injuries were significantly associated with the fatal outcome of COVID-19 (Guo et al. 2020).

After 100 days of outbreak of COVID-19 (14^th^ May, 2020), India rank 11^th^ with 2% of the global total COVID-19 confirmed cases and moving at a faster pace to surpass China. The case fatality ratio of India (3.2) is relatively lower but the number of testing per million population is low compared to many other countries. With a population of nearly 1.35 billion, India is at high risk of community transmission of COVID-19. The densely populated cities, with higher proportion of slum population coupled with poor health infrastructure, low income level and greater dependence on informal activities would raise the potential spread of COVID-19 pandemic. Projection suggests that the number of cases in India is reduced due to lockdown measures (Dwivedi et al. 2020) and may rise with ease of movement. The estimated cases are very low owing to low intensity of testing (Goli & James 2020).

Despite nationwide lock down and several effective measures for over two months in India, the number of COVID-19 confirmed cases was close to 170,000 mark with over 4,700 deaths as of 29^th^ May 2020. The disease has reached the stage of community infection in the states of Maharashtra, Gujarat, Delhi and Madhya Pradesh and no attempt has been made to understand the pattern of mortality associated with COVID-19 in India. The aim of this paper is to examine the possible age pattern of mortality under likely event of community infection of COVID-19 in India.

## Data and Methods

Data on COVID-19 at national and global level is made available in a seamless manner than any other disease. A number of key variables such as number of infection, deaths and recoveries were available for smallest geographic unit such as district to country level on daily basis. We have used data from the covid19india.org (https://www.covid19india.org/), worldmeter (https://www.worldometers.info/), the Sample Registration System (SRS) (ORGI, 2019) and the population projection of expert committee (MoHFW, 2019).

The covid19india.org provides micro data of patients on various domain such as demographics, death and recovered, testing number, travel history etc. (https://api.covid19india.org/csv). We have used raw data (patients’ data) and death and recovered data, as of 15^th^ May, 2020 (data updated till 9^th^ May). The raw data file provides selected variables such as detected district, state and city, age and sex, current status (hospitalised, recovered and death), nationality and status change date. While data on detected state, nationality and current status are available for most of the cases, data are largely missing for other variables. As of 9^th^ May 2020, there were 62,864 confirmed cases of which 19,301 were recovered and 2,100 were deceased. The age data was available for 7,191 hospitalised and recovered cases and 511 death cases. Besides, data from SRS abridged life table 2013-17 is used for the analyses (ORGI, 2019). We have used the SRS death rate to estimate deaths without COVID-19 since COVID-19 infection started in 2020. The expert committee on population projection of India 2011 has been used in estimation (MoHFW, 2019).

## Methods

Descriptive statistics, case-fatality ratio, lag case fatality ratio, estimation of longevity, years of Potential life lost (YPLL) and disability adjusted life years (DALY) are used in analyses. The set of assumptions used in estimation procedure are detailed below.

### Assumptions on Mortality without COVID-19 and with COVID-19

1. All deaths occurring due to COVID-19 are additional deaths in the population that could have been avoided in absence of COVID-19 cases in India
2. The age pattern of mortality obtained from SRS, 2013-17 was related to deaths without COVID-19 and assumed to remain same for all diseases other than COVID-19 for 2020.
3. Missing age data on hospitalisation and deaths have similar distribution as that of reported age
4. Effectiveness of medical treatment of COVID-19 will remain same till the end of the year 2020.

Age data on COVID-19 attributable mortality was available for 24% of all deceased (511) and that of hospitalised and recovery cases were available for 12% of total confirmed cases (7191). We have distributed the total COVID-19 attributed death cases in each of the five-year age group in keeping with the distribution of death for which age data is known. The official population projection of 2020, the age distribution of 2021 (2020 is not available) and the age specific death rate of SRS is used to arrive at expected number of deaths without COVID-19 (MoHFW 2019). The total number of deaths with COVID-19 is sum of deaths with and without COVID-19. The age specific death rate (ASDR) was estimated from all deaths including COVID-19 deaths and the key input in construction of abridge life table. The abridged life table is used to estimate the life expectancy under each of the four scenarios including that of without COVID-19.

### Estimation of case fatality ratio and lag case fatality ratio

The case fatality ratio is defined as the cumulative number of confirmed cases and cumulative number of deaths. Though it is a simple indictor and easy to understand, it underestimates the true mortality in the population (Baud et al. 2020). The lag case fatality ratio is the ratio of cumulative number of deaths and cumulative number of confirmed cases 14 days ago.

### Estimation of Years of Potential Life Lost (YPLL)

The YPLL is estimated as follows (Gardner & Sanborn, 1990),

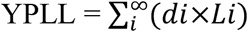

where,

*di* = Number of deaths in i^th^ age group

*Li* = Life expectancy at age i

### Estimation of Premature Years of Potential Life Lost (PYPLL)

The PYPLL is estimated as follows (Gardner & Sanborn, 1990),

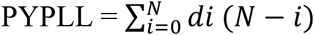

where, i= Age at death

*di* = Number of deaths at age i

N=Upper age limit (70 years)

### Estimation of Disability Adjusted Life Years (DALY)

DALY is a summary measure of population health, combining mortality and disability. The DALY is the sum of years of life lost (YLL) and years lived with disability (YLD).

The basic formula for YLL and YLD are:

YLL= L×N

YLD= I×DW×L

The formula used for estimation of DALY with discounting and age weighting are given below:

YLL = N/r (1- exp (-r*L))

where, N= number of deaths

L= Life expectancy at age of death

r= discount rate (e.g. 3% corresponds to a discount rate of 0.03)

YLD= (I ×DW × L (1- exp (-r*L))/ r

where, I= number of incidence/prevalence cases

DW= disability weight

L= duration of disability

r= discount rate

As COVID-19 is a novel disease, its disability weight is not available. Since COVID-19 is an infectious disease, we have used the weight of Infectious disease (severe) as proxy for COVID-19. Global Burden of Disease and World Health Organization have estimated this disability weight as 0.133 (Salomon et al. 2013). We have used duration of disability as 60 days because the patients of COVID-19 have been hospitalized for about 30 days and after discharged, quarantined for 14-28 days approximately.

## Results

### Rising Trends in COVID-19 cases

We begin the discussion by presenting the cumulative number of confirmed, deceased and recovered cases of India over a period of over two months (Fig 1). The exponential curve suggests that the average growth rate of 11% for total confirmed cases and 12% for deceased cases. With the current growth rate, the doubling time is estimated at 6.5 days for confirmed cases, 5 days for recovered and 5.5 days for death cases.

**Fig 1:**
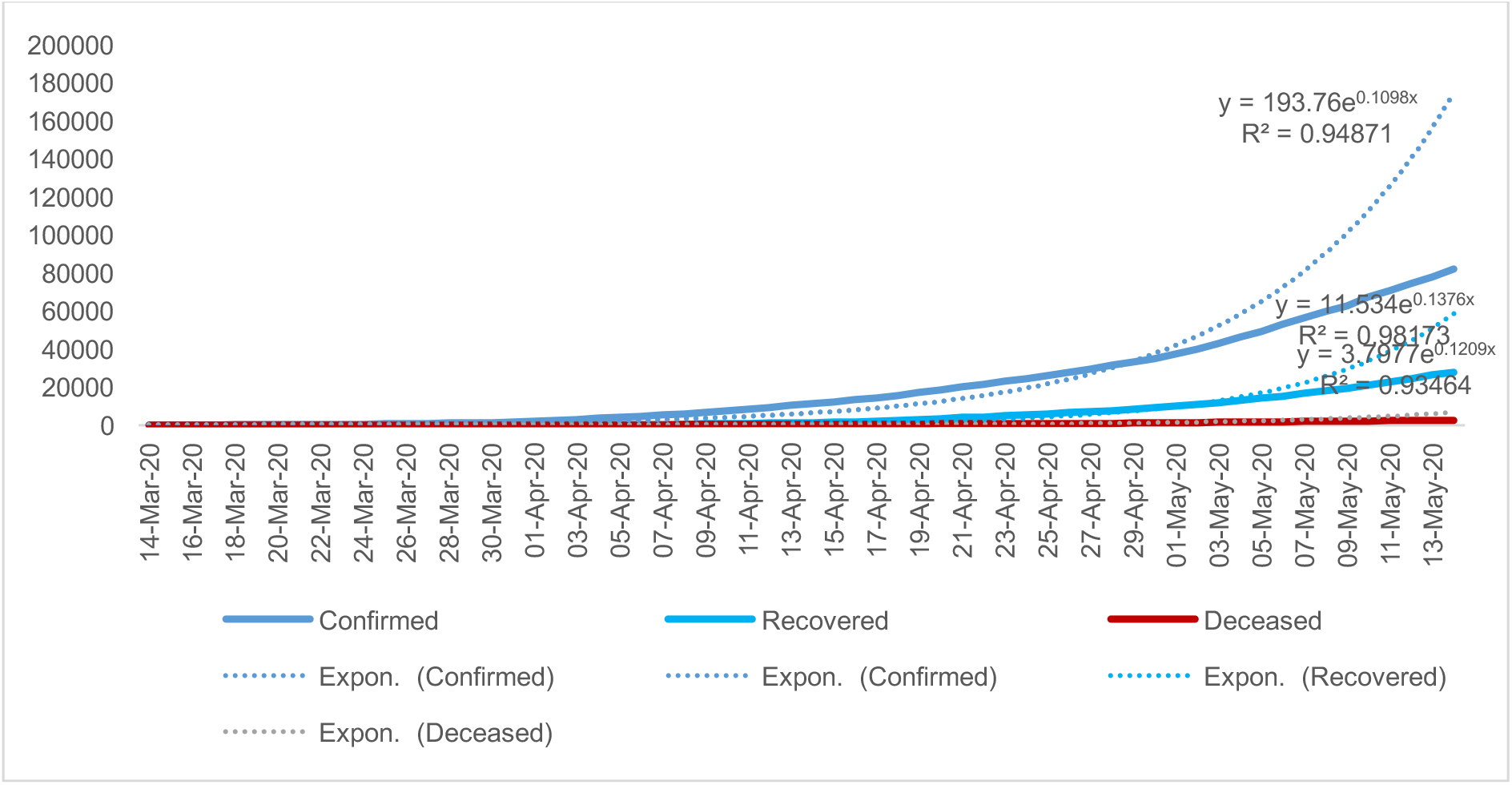
Trends in cumulative number of confirmed cases, recovered and death till 13^th^ May, 2020.

### Case Fatality Ratio and Lagged Case Fatality Ratio

Fig 2 presents trends in case fatality rate and lagged case fatality rate (LCFR) (with 14 days lag) in India on daily basis for over 47 days. There are at least two limitations of the CFR. First, even if there is no further new infection, there will be additional deaths from hospitalised or infected cases. Second, deaths occurring due to COVID-19 does not relate to infection on same day. The maximum incubation period of COVID-19 infection is 14 days and hence a refined measure that relate the deaths to number of infection 14 days prior to death. The LCFR was fluctuating in early days of infection but remained stable in last one month. From the graph, we infer that 1) the mortality level due to COVID-19 is at least twice higher than CFR 2) The LCFR has declined in initial period but remains stable in recent days.

**Fig 2:**
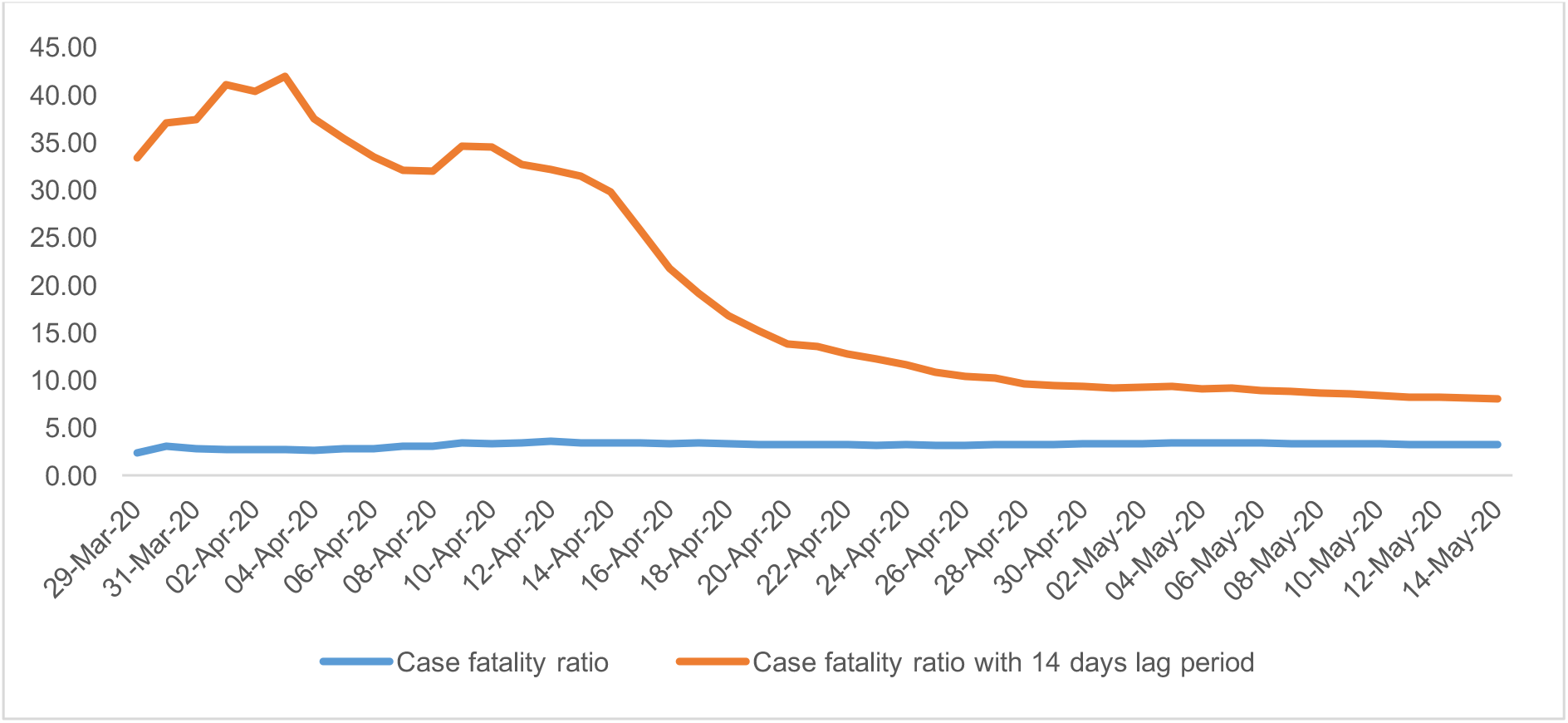
Trends in case fatality ratio and case fatality ratio with 14 days’ lag period in India, 29^th^ March-14^th^ May, 2020.

Table 1 presents the cumulative number of cases, death cases, the CFR and LCFR for selected states of India. Among these states, the LCFR was lowest in Kerala followed by Bihar and highest in West Bengal (29.7) followed by Gujarat (14.4) and Maharashtra (10.3). The LCFR for India is at least twice higher than CFR suggesting that the mortality due to COVID-19 is higher than that is conveyed by CFR.

**Table 1:** **Cumulative number of confirmed cases, deaths, case fatality ratio and case-fatality ratio with 14 days’ lag period in selected states of India during 30**^**th**^ **Jan-14**^**th**^ **May, 2020**

| States | Cumulative No of Total Cases | Cumulative No of death cases | Case Fatality Ratio (CFR) | Cumulative Number of cases with 14 days lag period | Lag Case Fatality Ratio (LCFR) |
| --- | --- | --- | --- | --- | --- |
| Kerala | 561 | 4 | 0.71 | 496 | 0.81 |
| Bihar | 999 | 7 | 0.70 | 403 | 1.74 |
| Tamil Nadu | 9674 | 66 | 0.68 | 2162 | 3.05 |
| Delhi | 8470 | 115 | 1.36 | 3439 | 3.34 |
| Telangana | 1414 | 34 | 2.40 | 1016 | 3.35 |
| Haryana | 818 | 11 | 1.34 | 311 | 3.54 |
| Andhra Pradesh | 2205 | 48 | 2.18 | 1332 | 3.60 |
| Uttar Pradesh | 3902 | 88 | 2.26 | 2134 | 4.12 |
| Rajasthan | 4534 | 125 | 2.76 | 2440 | 5.12 |
| Karnataka | 987 | 35 | 3.55 | 535 | 6.54 |
| **India** | **82047** | **2648** | **3.23** | **33065** | **8.01** |
| Madhya Pradesh | 4426 | 238 | 5.38 | 2560 | 9.30 |
| Maharashtra | 27524 | 1018 | 3.70 | 9915 | 10.27 |
| Gujarat | 9592 | 586 | 6.11 | 4082 | 14.36 |
| West Bengal | 2377 | 215 | 9.05 | 725 | 29.66 |

Table 2 presents the actual number of confirmed cases, adjusted number of cases (moving average) and the case fatality ratio of COVID-19 by age group in India. Total number of adjusted cases in India are distributed as per the distribution of total cases for which age is known and total number of confirmed cases. Findings suggest that about 60% of COVID-19 cases are in the age group of 30-64. About 2.3% deaths are under 15 years, 12.8% are in 15-44 age group, 48.2% deaths are in 45-64 age group and 36.8% are in 65+ age group. A major share of the COVID-19 deaths is in the working age group. The age specific case fatality ratio increases sharply from age 45 years and above.

**Table 2:** **Actual number of confirmed cases, adjusted number of confirmed cases, hospitalised, death cases and the case-fatality ratio by age due to COVID-19 only, India, 2020**

| Actual Number of Cases with given age |  |  |  |  | Adjusted Number of Cases |  |  |  |  |  |  |
| --- | --- | --- | --- | --- | --- | --- | --- | --- | --- | --- | --- |
| Age Group | Hospitalised and Recovered (Col 1) | Death (col 2) | Total number of Cases (col 3) | Distribution of known hospitalised and death data (col 4) | Moving average-Hospitalisation (col 5) | Smoothed cases - Hospitalisation (col 6) | Moving average-Death (col 7) | Smoothed cases - Death (col 8) | Percent distribution of COVID-19 deaths (col 9) | Total number of adjusted cases (all) (col 10) | Case Fatality Ratio (col 11) |
| 0-4 | 458 | 7 | 465 | 6.04 | 458 | 453 | 7 | 7 | 1.37 | 3795 | 1.50 |
| 5-9 | 176 | 0 | 176 | 2.29 | 272 | 269 | 3 | 3 | 0.59 | 1437 | 1.70 |
| 10-14 | 183 | 2 | 185 | 2.40 | 204 | 202 | 2 | 2 | 0.33 | 1510 | 0.90 |
| 15-19 | 253 | 3 | 256 | 3.32 | 348 | 344 | 4 | 4 | 0.72 | 2089 | 1.43 |
| 20-24 | 607 | 6 | 613 | 7.96 | 576 | 569 | 5 | 5 | 0.91 | 5003 | 0.76 |
| 25-29 | 867 | 5 | 872 | 11.32 | 843 | 834 | 6 | 6 | 1.17 | 7119 | 0.69 |
| 30-34 | 1054 | 7 | 1061 | 13.78 | 917 | 907 | 9 | 9 | 1.76 | 8662 | 0.85 |
| 35-39 | 829 | 15 | 844 | 10.96 | 883 | 873 | 15 | 15 | 2.86 | 6891 | 1.73 |
| 40-44 | 764 | 22 | 786 | 10.21 | 737 | 729 | 27 | 27 | 5.33 | 6415 | 3.47 |
| 45-49 | 618 | 45 | 663 | 8.61 | 609 | 603 | 43 | 43 | 8.32 | 5411 | 6.41 |
| 50-54 | 446 | 61 | 507 | 6.58 | 484 | 478 | 59 | 59 | 11.51 | 4138 | 11.60 |
| 55-59 | 387 | 71 | 458 | 5.95 | 360 | 356 | 67 | 67 | 13.13 | 3738 | 14.65 |
| 60-64 | 246 | 70 | 316 | 4.10 | 266 | 263 | 78 | 78 | 15.21 | 2579 | 24.60 |
| 65-69 | 165 | 93 | 258 | 3.35 | 160 | 158 | 70 | 69 | 13.59 | 2106 | 26.91 |
| 70-74 | 68 | 46 | 114 | 1.48 | 91 | 90 | 56 | 56 | 10.99 | 930 | 49.25 |
| 75-79 | 39 | 30 | 69 | 0.90 | 35 | 34 | 35 | 35 | 6.76 | 563 | 50.08 |
| 80+ | 30 | 28 | 58 | 0.75 | 30 | 30 | 28 | 28 | 5.46 | 473 | 48.12 |
| **Total** | **7191** | **511** | **7702** | **100.00** | **7271** | **7191** | **513** | **511** | **100.00** | **62862** |  |

Fig 3(a) presents the histogram on age of COVID-19 recovery among the hospitalised patients and Fig 3(b) presents that of deaths in India. The median age of hospitalisation cases was 34 years compared to 61 years of deceased. About one-fourth of deceased were below 70 years and that of hospitalised cases were below 46 years.

**Fig 3(a):**
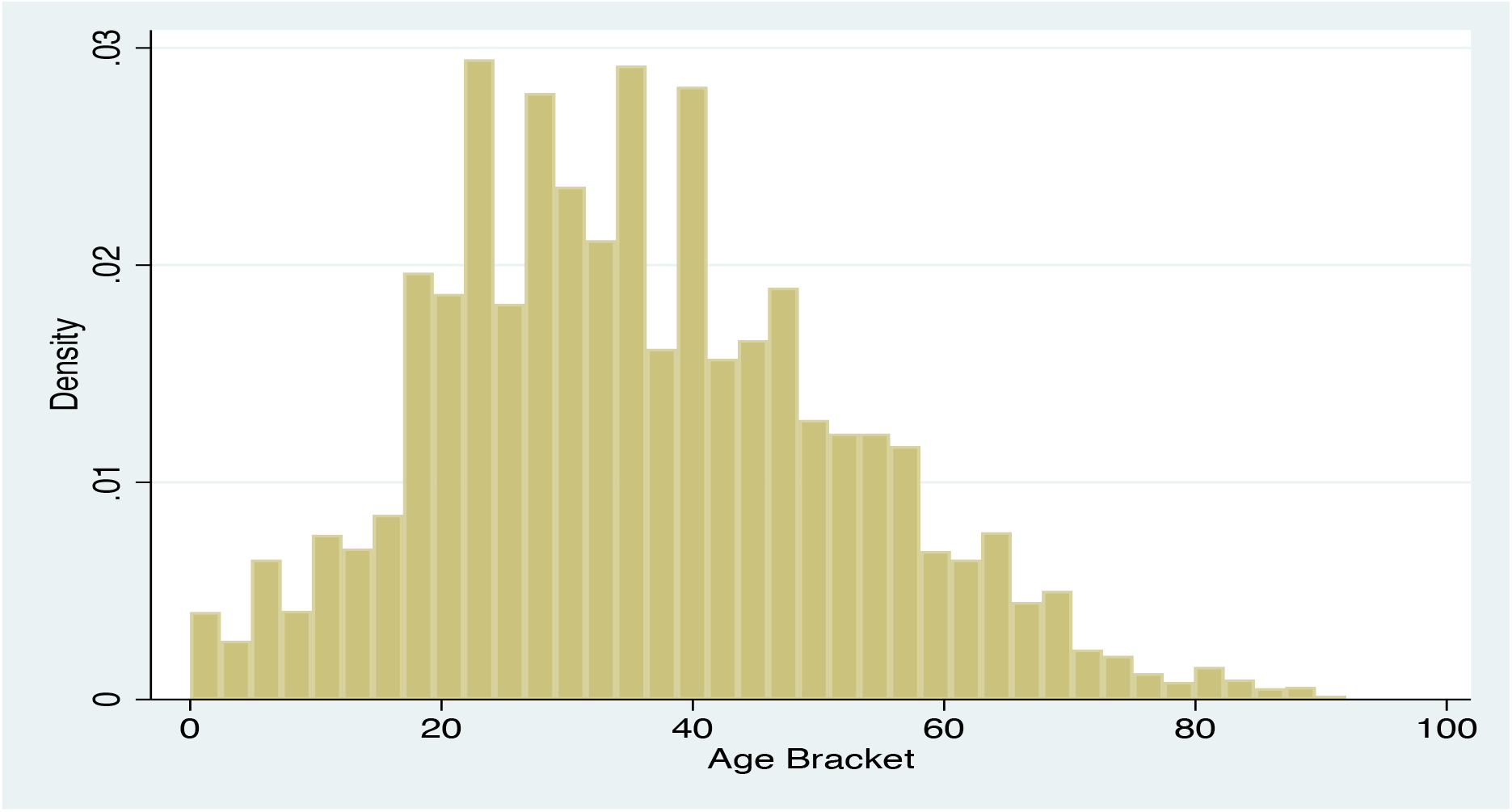
Histogram of hospitalised and recovered COVID-19 cases in India as of 9^th^ May, 2020.

**Fig S3(b):**
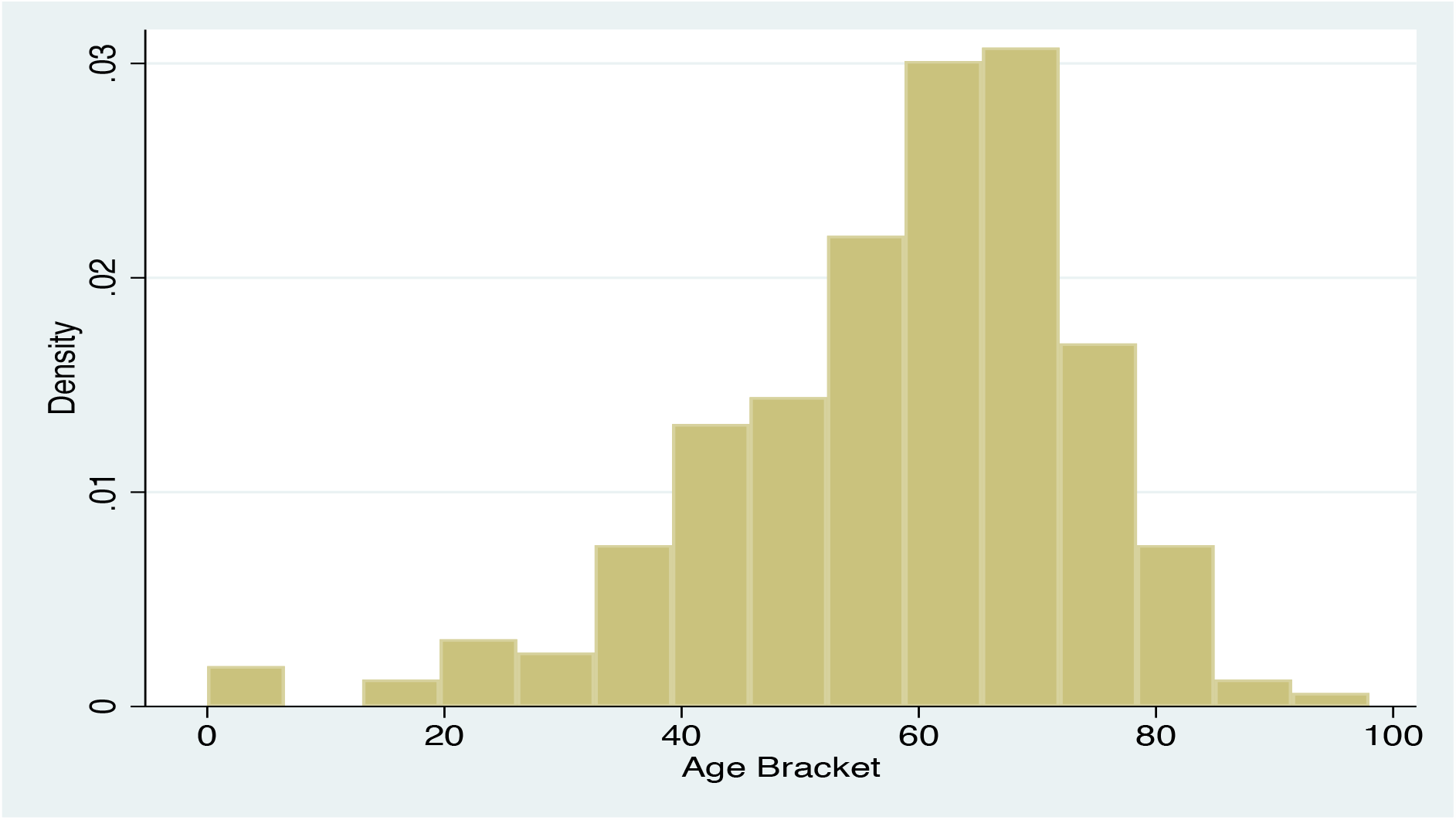
Histogram of deceased COVID-19 cases in India as of 9^th^ May, 2020.

**Fig 4:**
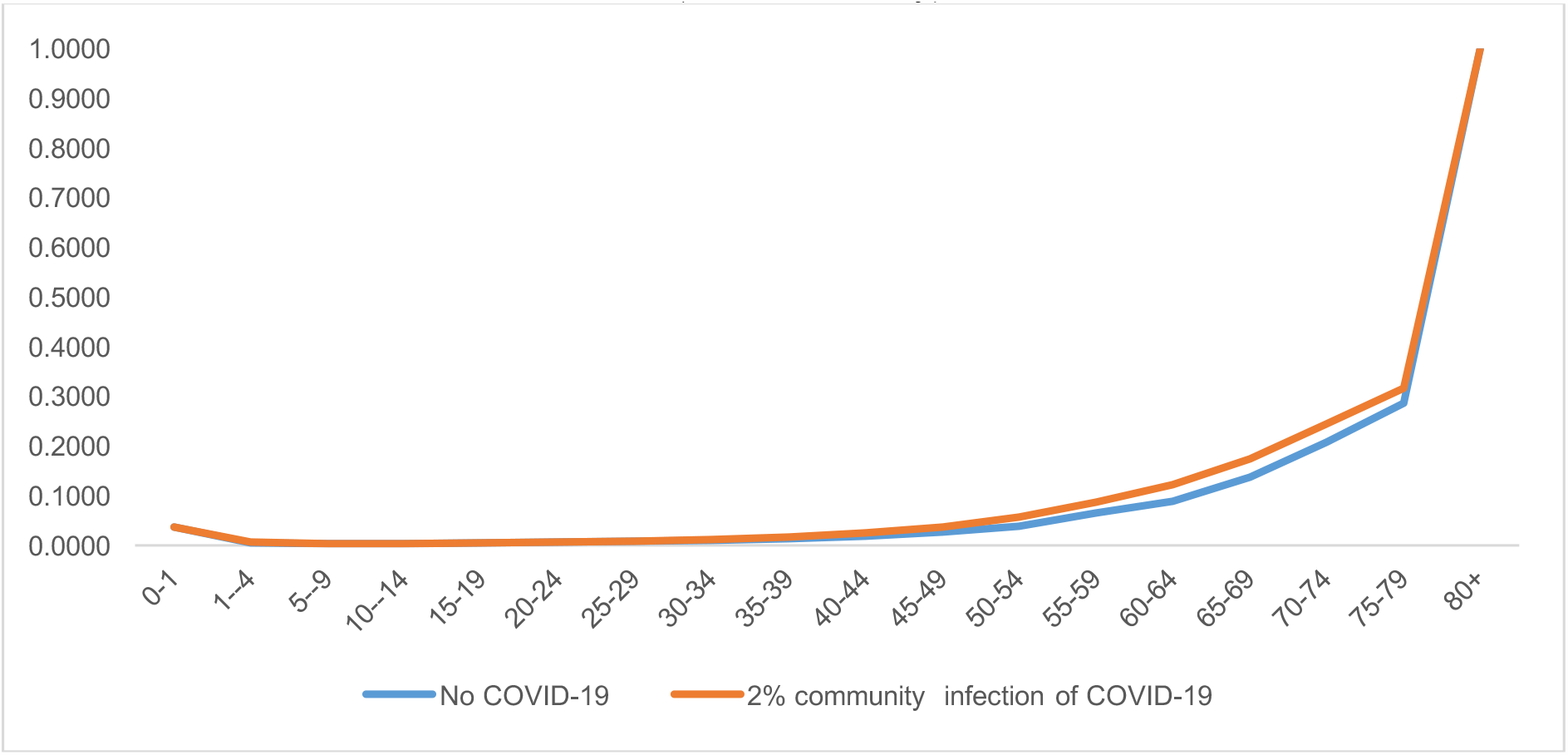
Life table probability of death without COVID-19 and with 2% community infection of COVID-19 in India, 2020 (as of 14^th^ May)

**Fig 5:**
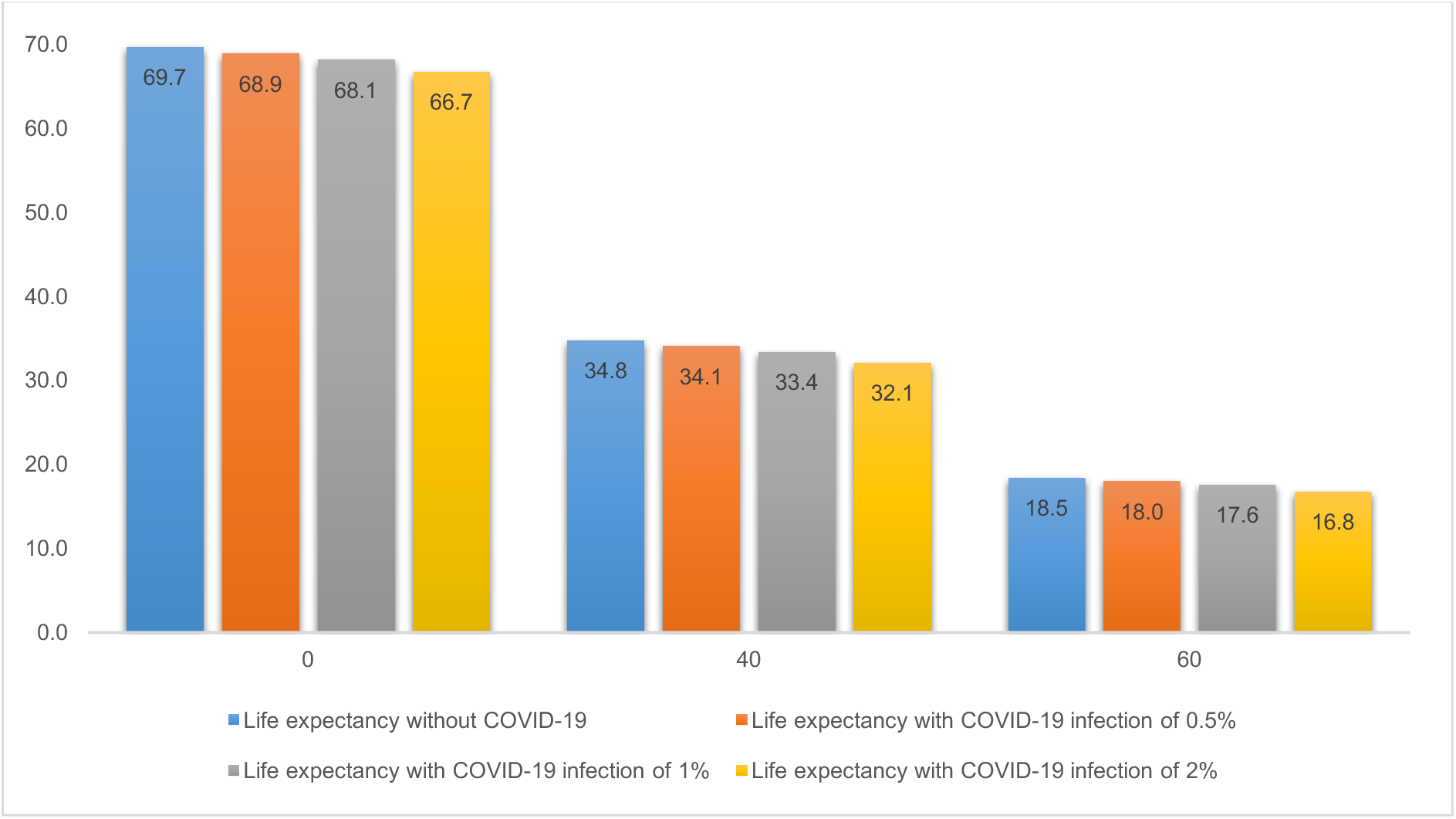
Life expectancy at exact age (0,40 and 60) by varying level of community infections of COVID-19 in India, 2020.

### Probability of Dying with and without COVID-19

Under this hypothetical situation, the age pattern of mortality clearly suggests that the COVID-19 is likely to affect the adults 45-75, majority in working age group. Beyond age 60, the probability of death due to COVID-19 would be at least four times higher among elderly compared with non-elderly. The probability of dying in 15-60 age group would increase from 0.17 without COVID-19 to 0.19 with 0.5% community infection, 0.20% with 1% community infection and 0.23 with 2% community infection.

### Life expectancy with and without COVID-19

Table 3 provides life expectancy and reduction in life expectancy at each age under three scenarios. It may be noticed that the life expectancy will be reduced in most of the age group under each scenario. If the community infection reaches at 0.5% by the end of the year, the life expectancy is likely to reduce by 0.8 years from 2013-17 level. If the infection is at 1% level, the life expectancy would reduce by 1.5 years while an infection rate of 2% would reduce the life expectancy by 3.0 years.

**Table 3:** **Life expectancy without COVID-19 and with COVID-19 under alternative scenario in India, 2020**

| Age group | Life expectancy without COVID-19 | Life expectancy with COVID-19 infection of 0.5% | Life expectancy with COVID-19 infection of 1% | Life expectancy with COVID-19 infection of 2% | Reduction in life expectancy with COVID-19 infection of 0.5% | Reduction in life expectancy with COVID-19 infection of 1% | Reduction in life expectancy with COVID-19 infection of 2% | Reduction in life expectancy (%) with COVID-19 infection of 0.5% | Reduction in life expectancy (%) with COVID-19 infection of 1% | Reduction in life expectancy (%) with COVID-19 infection of 2% |
| --- | --- | --- | --- | --- | --- | --- | --- | --- | --- | --- |
| 0-1 | 69.7 | 68.9 | 68.1 | 66.7 | 0.8 | 1.5 | 3.0 | 1.13 | 2.21 | 4.26 |
| 1-4 | 71.3 | 70.5 | 69.7 | 68.2 | 0.8 | 1.6 | 3.1 | 1.14 | 2.24 | 4.30 |
| 5-9 | 67.6 | 66.8 | 66.1 | 64.7 | 0.8 | 1.5 | 2.9 | 1.14 | 2.24 | 4.31 |
| 10-14 | 62.8 | 62.1 | 61.3 | 59.9 | 0.8 | 1.5 | 2.9 | 1.22 | 2.39 | 4.60 |
| 15-19 | 58.0 | 57.2 | 56.5 | 55.1 | 0.8 | 1.5 | 2.9 | 1.32 | 2.58 | 4.96 |
| 20-24 | 53.2 | 52.5 | 51.8 | 50.4 | 0.8 | 1.5 | 2.9 | 1.43 | 2.79 | 5.37 |
| 25-29 | 48.6 | 47.8 | 47.1 | 45.7 | 0.8 | 1.5 | 2.8 | 1.55 | 3.04 | 5.84 |
| 30-34 | 43.9 | 43.2 | 42.4 | 41.1 | 0.7 | 1.5 | 2.8 | 1.70 | 3.34 | 6.41 |
| 35-39 | 39.3 | 38.6 | 37.9 | 36.5 | 0.7 | 1.4 | 2.8 | 1.88 | 3.68 | 7.05 |
| 40-44 | 34.8 | 34.1 | 33.4 | 32.1 | 0.7 | 1.4 | 2.7 | 2.07 | 4.05 | 7.76 |
| 45-49 | 30.4 | 29.7 | 29.1 | 27.9 | 0.7 | 1.3 | 2.6 | 2.23 | 4.38 | 8.40 |
| 50-54 | 26.2 | 25.5 | 24.9 | 23.8 | 0.6 | 1.2 | 2.3 | 2.37 | 4.63 | 8.89 |
| 55-59 | 22.1 | 21.6 | 21.1 | 20.1 | 0.5 | 1.0 | 2.0 | 2.42 | 4.74 | 9.11 |
| 60-64 | 18.5 | 18.0 | 17.6 | 16.8 | 0.4 | 0.9 | 1.7 | 2.40 | 4.71 | 9.08 |
| 65-69 | 15.0 | 14.7 | 14.3 | 13.7 | 0.3 | 0.6 | 1.2 | 2.19 | 4.30 | 8.31 |
| 70-74 | 12.0 | 11.7 | 11.5 | 11.1 | 0.2 | 0.4 | 0.9 | 1.88 | 3.70 | 7.18 |
| 75-79 | 9.4 | 9.3 | 9.1 | 8.9 | 0.1 | 0.3 | 0.6 | 1.55 | 3.05 | 5.93 |
| 80+ | 7.2 | 7.1 | 7.0 | 6.8 | 0.1 | 0.2 | 0.4 | 1.41 | 2.79 | 5.43 |

\*Note: LCFR: Lag Case-fatality ratio

Table 4 presents the estimated YPLL in the absence of COVID-19 and at varying level of COVID-19 infection. Without any COVID, YPLL was 399 million from all disease with majority of share among children (55% among 0-1 age group), 15% among 45-64 years and 14% among 65 and above. YPLL was 12 million with 0.5% infection of COVID-19. However, the age pattern of YPLL due to COVID-19 suggests that it is largely affecting the working population, aged 45-64 years. The pattern remains similar even at higher level of community infection.

**Table 4:** **Age pattern of years of potential life lost (YPLL) under varying scenarios of COVID-19 infection in India, 2020**

| Age Group | No COVID-19 |  | COVID infection at 0.5% |  | COVID infection at 1 % |  | COVID infection at 2% |  |
| --- | --- | --- | --- | --- | --- | --- | --- | --- |
|  | YPLL in millions | Share of YPLL | YPLL in millions | Share of YPLL | YPLL in millions | Share of YPLL | YPLL in millions | Share of YPLL |
| 0-1 | 220.03 | 55.02 | 0.21 | 1.73 | 0.41 | 1.73 | 0.83 | 1.76 |
| 1-4 | 2.20 | 0.55 | 0.32 | 2.63 | 0.63 | 2.66 | 1.27 | 2.70 |
| 5-9 | 4.67 | 1.17 | 0.21 | 1.78 | 0.43 | 1.80 | 0.86 | 1.83 |
| 10-14 | 4.44 | 1.11 | 0.11 | 0.92 | 0.22 | 0.93 | 0.44 | 0.94 |
| 15-19 | 6.44 | 1.61 | 0.23 | 1.86 | 0.45 | 1.88 | 0.90 | 1.91 |
| 20-24 | 8.14 | 2.04 | 0.26 | 2.18 | 0.53 | 2.19 | 1.05 | 2.22 |
| 25-29 | 8.69 | 2.17 | 0.31 | 2.55 | 0.62 | 2.56 | 1.23 | 2.59 |
| 30-34 | 9.16 | 2.29 | 0.42 | 3.45 | 0.84 | 3.46 | 1.67 | 3.49 |
| 35-39 | 10.50 | 2.63 | 0.61 | 5.02 | 1.22 | 5.03 | 2.44 | 5.05 |
| 40-44 | 11.65 | 2.91 | 1.01 | 8.27 | 2.01 | 8.27 | 4.03 | 8.28 |
| 45-49 | 12.65 | 3.16 | 1.37 | 11.27 | 2.75 | 11.25 | 5.50 | 11.22 |
| 50-54 | 14.28 | 3.57 | 1.63 | 13.38 | 3.27 | 13.34 | 6.53 | 13.26 |
| 55-59 | 16.74 | 4.19 | 1.58 | 12.90 | 3.15 | 12.86 | 6.30 | 12.76 |
| 60-64 | 15.19 | 3.80 | 1.52 | 12.48 | 3.05 | 12.44 | 6.10 | 12.35 |
| 65-69 | 14.88 | 3.72 | 1.11 | 9.07 | 2.21 | 9.06 | 4.42 | 9.03 |
| 70-74 | 14.24 | 3.56 | 0.71 | 5.87 | 1.43 | 5.89 | 2.85 | 5.90 |
| 75-79 | 11.09 | 2.77 | 0.35 | 2.86 | 0.69 | 2.88 | 1.39 | 2.90 |
| 80+ | 14.91 | 3.73 | 0.21 | 1.77 | 0.43 | 1.78 | 0.86 | 1.80 |
| **Total** | **399.90** | **100.00** | **12.17** | **100.00** | **24.34** | **100.00** | **48.68** | **100.00** |
| 0-44 | 285.92 | 71.50 | 3.68 | 30.25 | 7.36 | 30.25 | 14.73 | 30.25 |
| 45-64 | 58.85 | 14.72 | 6.11 | 50.19 | 12.22 | 50.19 | 24.43 | 50.19 |
| 65+ | 55.12 | 13.78 | 2.38 | 19.56 | 4.76 | 19.56 | 9.52 | 19.56 |
| **Rate of YPLL** | **295** |  | **9** |  | **18** |  | **36** |  |

Table 5 shows the estimated DALY with discounting and age weighting in the three scenarios of COVID-19 infection. The estimated DALY is 6.2 per thousand population with 0.5% infection of COVID-19 which would increase to 12.3 and 24.6 per thousand population with 1% and 2% infection, respectively. Age-group 45-64 makes the highest contribution to the overall DALY. As COVID-19 is a short term but fatal disease, the YLL component of DALY is higher than the YLD.

**Table 5:** **Age pattern of Year of life lost (YLL), Years lived with Disability (YLD) and Disability adjusted life years (DALY) (in per 1000 population) in alternative scenarios of COVID-19 infection in India, 2020**

| Age Group | YLL per 1000 population |  |  | YLD per 1000 population |  |  | DALY per 1000 population |  |  |
| --- | --- | --- | --- | --- | --- | --- | --- | --- | --- |
|  | COVID-19 infection at 0.5% | COVID-19 infection at 1% | COVID-19 infection at 2% | COVID-19 infection at 0.5% | COVID-19 infection at 1% | COVID-19 infection at 2% | COVID-19 infection at 0.5% | COVID-19 infection at 1% | COVID-19 infection at 2% |
| 0-1 | 1.03 | 2.05 | 4.10 | 0.01 | 0.02 | 0.03 | 1.03 | 2.07 | 4.14 |
| 1-4 | 4.66 | 9.32 | 18.65 | 0.04 | 0.08 | 0.16 | 4.70 | 9.40 | 18.80 |
| 5-9 | 0.80 | 1.60 | 3.19 | 0.01 | 0.01 | 0.02 | 0.80 | 1.61 | 3.22 |
| 10-14 | 0.42 | 0.85 | 1.69 | 0.01 | 0.01 | 0.02 | 0.43 | 0.86 | 1.72 |
| 15-19 | 0.87 | 1.73 | 3.46 | 0.01 | 0.02 | 0.03 | 0.87 | 1.75 | 3.49 |
| 20-24 | 1.03 | 2.06 | 4.12 | 0.02 | 0.04 | 0.08 | 1.05 | 2.10 | 4.20 |
| 25-29 | 1.36 | 2.72 | 5.45 | 0.03 | 0.06 | 0.11 | 1.39 | 2.78 | 5.56 |
| 30-34 | 2.12 | 4.24 | 8.47 | 0.04 | 0.08 | 0.15 | 2.16 | 4.31 | 8.62 |
| 35-39 | 3.62 | 7.24 | 14.49 | 0.03 | 0.07 | 0.13 | 3.66 | 7.31 | 14.62 |
| 40-44 | 7.09 | 14.19 | 28.38 | 0.03 | 0.07 | 0.14 | 7.13 | 14.26 | 28.52 |
| 45-49 | 11.27 | 22.54 | 45.08 | 0.03 | 0.06 | 0.13 | 11.30 | 22.60 | 45.21 |
| 50-54 | 16.38 | 32.76 | 65.51 | 0.03 | 0.06 | 0.11 | 16.41 | 32.81 | 65.63 |
| 55-59 | 20.24 | 40.47 | 80.95 | 0.03 | 0.06 | 0.13 | 20.27 | 40.54 | 81.08 |
| 60-64 | 26.17 | 52.33 | 104.66 | 0.03 | 0.06 | 0.11 | 26.19 | 52.39 | 104.77 |
| 65-69 | 26.28 | 52.57 | 105.13 | 0.03 | 0.06 | 0.12 | 26.31 | 52.63 | 105.25 |
| 70-74 | 23.29 | 46.57 | 93.14 | 0.02 | 0.03 | 0.07 | 23.30 | 46.61 | 93.21 |
| 75-79 | 17.12 | 34.25 | 68.49 | 0.02 | 0.03 | 0.06 | 17.14 | 34.28 | 68.55 |
| 80+ | 12.90 | 25.80 | 51.60 | 0.02 | 0.03 | 0.06 | 12.92 | 25.83 | 51.66 |
| **Total** | **6.14** | **12.27** | **24.54** | **0.02** | **0.04** | **0.09** | **6.16** | **12.32** | **24.63** |

## Discussion and Conclusion

This is the first ever systematic attempt at assessing the likely effect of COVID-19 spread on mortality and longevity in India. It compares a situation with varying levels of community infection of COVID-19. The followings are the salient findings of the study. First, India fares better than global average and many other worst affected countries as regard fatality rate, recovery rate and test detection ratio. However, it the mortality due to COVID-19 is measured using case fatality rate with a lag of 14 days, it is twice as high as conventional CFR. The inter-state variation in LCFR was large with West Bengal, Maharashtra and Gujarat being three worst performing states. Second, the community spread of COVID-19 to be restricted at 0.5% of the population in India would result in 0.8 years compression of the life expectancy at birth. Third, the impact of COVID-19 on premature mortality, particularly among working adults in 45-64 age group is very high. Majority of the deaths being in the ages 0f 45-64, the contribution to YPLL and DALY is likely to be the most by 45-64 years age group. Lastly, this exercise is an instance of effective provisioning of the patient level and aggregated CVID-19 data for research and policy formulation in India. However, care must be taken to sensitise the officials/health workers in providing complete information on key variables to facilitate its scientific treatment.

We provide some explanation and suggestion in support of the findings. The low fatality rate and high test-detection ratio are primarily due to effective measured adopted by Govt. of India in controlling the community spread of the disease. The lockdown measures of about two months is the most proactive step taken by the national government in curbing the epidemic. But the lockdown cannot be continued indefinitely given its economic cost as observed in terms of reassessment of growth. The pace of new infection and deaths due to COVID-19 is soaring every day. It is believed to have reached the community level in the states of Maharashtra, Gujarat and Delhi. The situation is alarming in Maharashtra and particularly in Mumbai city. Many of the COVID-19 patients are not getting hospital in civic run hospitals and families of positive patients are not getting tested anymore. However, it is interesting to note the control of COVID-19 in Kerala and the best practice adopted in the Kerala may well be practised in worst affected states of India. Efforts to minimise the loss of human life needs preparedness of health facilities to cater to severe patients apart from imposing strict adherence of the ‘new normal’ (i.e. social distancing and hand hygiene). Identification of high-risk cases and arrangement of intensive care for such patients would help reducing premature mortality. While community testing in India does not seem viable, it is necessary to provide testing treatment options for this infection in every town and cities of the country.

The study suffers from following limitations. We believe that our estimates for elderly is underestimated as missing data is likely to be higher among elderly compared to younger population. Our assumption of all deaths due to COVID-19 would have been avoided if there were no COVID-19 cases may not always hold true and the assumption of SRS death rate of 2013-17 would prevail is illustrative. Despite these limitations, we believed that this study offers a reasonable understanding of the impact of COVID-19 on longevity in India. It has demonstrated the scope to replicate similar analyses elsewhere and guide the impact assessment of the epidemic in the national and global scene.

### Research Ethics Approval

The study does not involve human participation. It used secondary data available in public domain and research ethics approval not required.

## Data Availability

https://api.covid19india.org/csv.

https://population.un.org/wpp/DataQuery/

https://www.worldometers.info/

